# Quantitative MRI and histopathology detect remyelination in inactive multiple sclerosis lesions

**DOI:** 10.1101/2022.08.01.22271457

**Authors:** Vanessa Wiggermann, Verena Endmayr, Enedino Hernández-Torres, Romana Höftberger, Gregor Kasprian, Simon Hametner, Alexander Rauscher

## Abstract

Magnetic resonance imaging (MRI) of focal or diffuse myelin damage or remyelination may provide important insights into disease progression and potential treatment efficacy in multiple sclerosis (MS). We performed post-mortem MRI and histopathological myelin measurements in seven progressive MS cases to evaluate the ability of three myelin-sensitive MRI scans to distinguish different stages of MS pathology, particularly chronic demyelinated and remyelinated lesions.

At 3 Tesla, we acquired two different myelin water imaging (MWI) scans and magnetization transfer ratio (MTR) data. Histopathology included histochemical stainings for myelin phospholipids (LFB) and iron as well as immunohistochemistry for myelin proteolipid protein (PLP), CD68 (phagocytosing microglia/macrophages) and BCAS1 (remyelinating oligodendrocytes). Mixed-effects modelling determined which histopathological metric best predicted MWF and MTR in normal appearing and diffusely abnormal white matter, slowly expanding, inactive, remyelinated and ischemic lesions.

Both MWI measures correlated well with each other and histology across regions, reflecting the different stages of MS pathology. MTR data showed a considerable influence of components other than myelin and a strong dependency on tissue storage duration. Both MRI and histology revealed increased myelin densities in inactive compared with slowly expanding lesions. Chronic inactive lesions harboured single scattered myelin fibres indicative of low-level remyelination. Mixed-effects modelling showed that smaller differences between white matter areas were linked to PLP densities and only to a smaller extent confounded by iron.

MWI reflects differences in myelin lipids and proteins across all levels of myelin densities encountered in MS, including low-level remyelination in chronic inactive lesions.

---

**M**ultiple sclerosis (MS) is a chronic inflammatory disease of the central nervous system (CNS), leading to the formation of focal areas of destruction, phagocytosis and digestion of CNS myelin sheaths. Remyelination may follow demyelination (1), but the proportion of remyelinated MS lesions varies greatly between individuals at autopsy (2). Fostering remyelination reduces axonal loss and may thus curtail MS progression (3), rendering it an important target of new therapeutic trials. Non-invasive myelin quantification by means of magnetic resonance imaging (MRI) will be a key component of such trials (4). Among the many MRI techniques for assessing myelin in vivo, multi-echo spin-echo T_2_ MWI (5–9) is considered one of the most myelin-specific (10–13) as it separately quantifies the MRI signal from water protons in the myelin phospholipid bilayers. The mobility of these protons is highly restricted, resulting in a faster T_2_ decay than water protons within axons, cell bodies and the extracellular space. The ratio of the short T_2_ signal component over the total water signal, termed myelin water fraction (MWF), is considered a surrogate measure of myelin content (5, 14).

Here, we compared three MRI-based myelin scans for the quantitation of myelin in seven progressive MS cases. MS lesions in various stages of deand remyelination, normal appearing and diffusely abnormal white matter were included and characterized with histopathology and MRI-based myelin measures: 3D multi-echo T_2_ gradient- and spin-echo (GraSE) MWI (15), 3D multi-echo spin-echo MWI (16, 17) and 3D MTR (18). Histopathology included Luxol Fast Blue (LFB) staining for phospholipids and diaminobenzidine-enhanced Turnbull blue (TBB) staining for iron. In addition, immunohistochemical stainings for proteolipid protein (PLP), one of the major myelin proteins, CD68 for phagocytosing microglia/macrophages and BCAS1 staining for remyelinating oligodendrocytes were obtained (19).

## Materials and Methods

### Post-mortem samples

Seven *post-mortem* brains of progressive MS patients (six with secondary-progressive MS, one with primary-progressive MS) were included. Whole brains had been fixed in 4% neutral-buffered formalin for three to four weeks at room temperature. Four cases from the archive at the Neuroimmunology department of the Center for Brain Research, Medical University of Vienna (MUV), Austria, had been fixed in formalin and stored for 9 to 14 years in 1%formalin diluted in phosphate-buffered saline (PBS) at 4^°^ C. Three cases derived from the Division of Neuropathology and Neurochemistry, MUV, Austria, were fixed and stored in formalin for up to 1 year. Basic epidemiological and clinical data are provided in Table 1.

**Table 1.** Demographic information of the *post-mortem* brain samples, including approximate time since fixation in years. To preserve anonymity, only age ranges are provided. Causes of death were unrelated to MS and included myocardial infarction, cardiovascular death and pancreatic cancer. Storage time refers to storage time in 1% paraformaldehyde / phosphate-buffered saline (SPMS: secondary progressive MS; PPMS: primary progressive MS; M: male; F: female)

|  | Age<br>w/i [yrs] | Sex | Clinical<br>course | Disease<br>dur. [yrs] | Storage<br>[yrs] |
| --- | --- | --- | --- | --- | --- |
| Case 1 | 31-35 | M | SPMS | 10.0 | 10.0 |
| Case 2 | 41-45 | M | SPMS | 11.4 | 14.0 |
| Case 3 | 46-50 | F | SPMS | 37.0 | 9.0 |
| Case 4 | 66-70 | M | PPMS | 7.3 | 12.0 |
| Case 5 | 61-65 | F | SPMS | 33 | < 1 |
| Case 6 | 51-55 | M | SPMS | 24 | < 1 |
| Case 7 | 56-60 | F | SPMS | 34 | < 1 |

### MRI data acquisition

For Cases 6 and 7, whole brains were available for MRI. Of all other cases, smaller tissue slabs were scanned. Their diameters ranged from 21.1 to 110.2 mm and their thickness from 4.2 to 23.8 mm. Scanning was performed on two identical 3T MRI systems (Philips Achieva, Best, The Netherlands) in 2015-2016; four cases at the University of British Columbia (UBC) and three cases at MUV. For all samples, we acquired 32-echo 3D-GraSE T_2_ data for MWI (TE/ΔTE/TR = 10/10/719 ms, reconstructed 0.9 *×* 0.9 *×* 1.5 mm^3^) (15). 3D-MTR images were acquired for all cases, except Case 6 (TE/TR = 3.7/84.6 ms, reconstructed 0.9 *×* 0.9 *×* 2 mm^3^, flip angle(exc/mtc) = 18^°^/520^°^, Δf = 1.1 kHz below Larmor frequency). Data acquisition at UBC (Cases 1-4) also included 3D multi-echo spin-echo imaging (CPMG – Carr-Purcell Meiboom-Gill, TE/ΔTE/TR = 10/10/1200 ms, reconstructed 0.9 *×* 0.9 *×* 3 mm^3^) (16, 17). T_2_-weighted images were acquired for registration purposes at TE/TR = 100 ms/2500 ms, 0.66*×*0.66*×*0.7 mm^3^. Acquisition parameters were minimally adapted to adjust for the different sample sizes. Only for Case 5, the 3D-GraSE sequence was substantially changed necessitated by the much larger field-of-view. The study and tissue scanning protocol were approved by the clinical research ethics boards of both sites (UBC: H15-011, MUV: EK Nr. 1491-2017).

### MRI data processing

Data processing and analysis was implemented in MATLAB 2017b. MWF was computed as previously described (5, 20). Briefly, T_2_ distributions were obtained using the extended phase graph algorithm (21) and regularized, non-negative least squares fitting. T_2_ distributions of all tissue samples in multiple white matter (WM) regions were consulted. Subsequently, the MWF was determined as the T_2_ signal fraction between 12 – 25 ms relative to all water (intra/extracellular water T_2_ range: 25 – 100 ms). MTR maps were calculated as the ratio of the signal difference between the on-resonance and off-resonance scan relative to the off-resonance scan (18).

### Tissue Preparation

After scanning, whole-brain cases were cut into 1 cm thick coronal double-hemispheric slices. All other tissue slabs were directly processed for histology. Samples were dehydrated (Miles Scientific™ autotechnicon device) and embedded in paraffin. Paraffin blocks were cut into 10 *μ*m thick serial sections with a tetrander microtome (R. Jung AG, Heidelberg). Sections were stained for hematoxylin and eosin (H&E) to assess general pathology, Luxol fast blue – periodic-acid Schiff (LFB-PAS) to visualize myelin and classify MS lesions, and diaminobenzidine (DAB)-enhanced TBB to detect iron (22). Iron is stored in myelin-forming oligodendrocytes, but also in myelin itself and is a known confounder to MWI and MTR (23–25). Immunohistochemistry for mouse monoclonal PLP (Serotec, MCA839G, dilution 1:1000) was performed by heat-steaming sections in ethylenediaminetetraacetic acid (EDTA, pH 9.0, 1h) for antigen retrieval. Unspecific antibody binding was blocked by incubation in 10% fetal calf serum (FCS) diluted in Dako wash buffer (DAKO, 20 min, room temperature). After overnight-incubation at 4^°^ C, a biotinylated donkey anti-mouse secondary antibody was applied. Sections were then incubated with avidin-conjugated horseradish peroxidase (Sigma Aldrich, dilution 1:500 in 10% FCS/DAKO, 1h, room temperature) and developed with the chromogen DAB, producing a brown PLP signal. For CD68 (Dako, M0814, Clone KP1, dilution 1:2000) and BCAS1 (Santa Cruz, sc-136342, dilution 1:4000), sections were steamed in TRIS buffer (pH 6.0, 1h). Primary antibodies were applied overnight at 4^°^ C. After application of an appropriate biotinylated secondary anti-mouse antibody, the Envision system was used according to the manufacturer’s protocol. Stainings were visualized by DAB development for 2 *×* 5 minutes, producing a brown reaction product. Stainings were scanned using an Agfa Duoscan^®^ device under standardized brightness conditions, at 800 pixels/inch, and saved as RGB TIFF.

### MRI-histology registration

Figure 1 provides an overview of the MRI-histology registration procedure. Histology images were downsampled by a factor of two along both dimensions. LFB-PAS and PLP sections were colour-deconvolved using ImageJ version 1.43r (A.C. Ruifrok, NIH, Bethesda, MD, USA) to extract the blue channel, i.e. the LFB dye, of the LFB-PAS double-stained sections and the red channel, i.e. the DAB dye, of the hematoxylin-counterstained PLP sections. Because mounting of the sections on glass slides can introduce small 2D distortions, TBB and PLP were registered to the LFB section using 2D FSL FLIRT (26) (default parameters, five degrees of freedom). Initial registrations utilized sections prior to colour deconvolution. Registration matrices were then applied to the lower-contrast, colour-deconvolved sections.

**Fig. 1.**
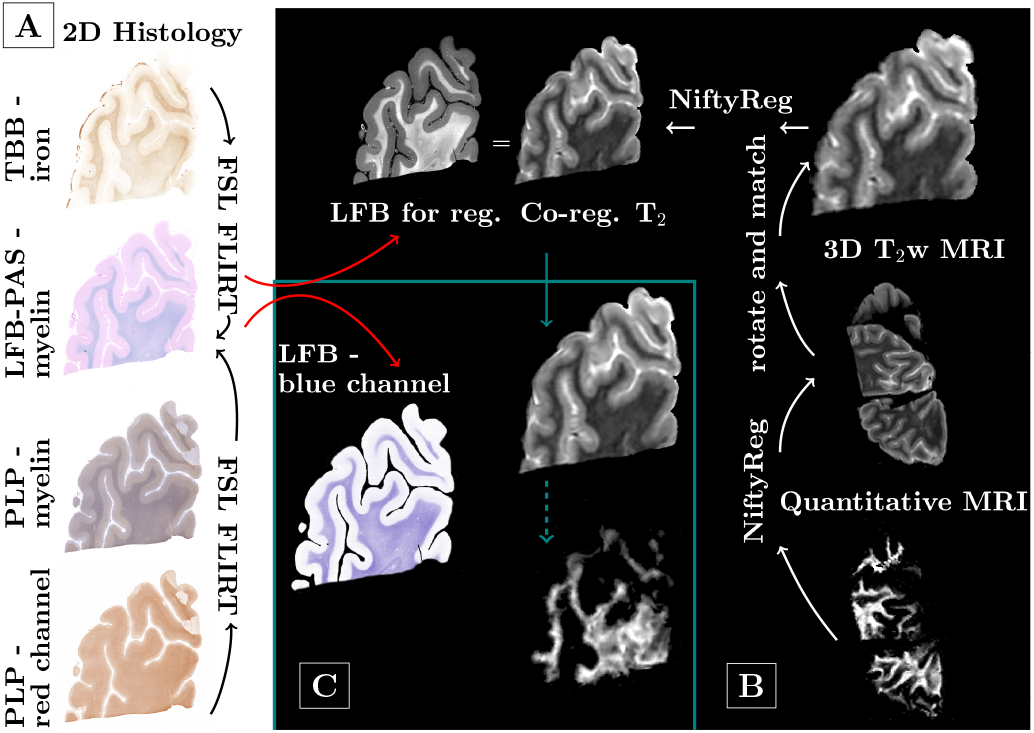
Overview of processing pipeline. Differently stained histology sections were aligned with 2D FSL FLIRT. To register all quantitative images, first a pipeline was established based on the 3D T2 scans, which were reoriented and resliced to match the histology. Registration between histology and MRI was performed using NiftyReg. Quantitative maps were then registered to the 3D T2 and the already established pipeline and registration matrices were applied.

Within-MRI and histology-MRI registrations were performed using NiftyReg’s aladin and f3d (default parameters) (27). T_2_-weighted MRIs were reoriented and sliced to match the histology section plane. To approximate the resolutions, reformatted images were up-sampled and histology images further down-sampled. Manual corrections, such as removal of tissue from unrelated sections, were performed for all cases individually. Lastly, registered images were up-sampled to match the histology resolution. MRI data were sent to the histology space to avoid altering the regions-of-interest (ROIs), which were manually defined on the histology sections in FSLview. For the quantitative analysis, all 3D MRIs were registered to the T_2_-weighted images. Linear registrations were performed using individual echoes of the quantitative data (default parameters). Subsequently, quantitative maps were transformed using NiftyReg’s resample and were then subjected to the same pipeline as the T_2_-weighted images, i.e. volume re-orientation, slice selection and mapping to histology.

### Manual ROI definition

ROIs were identified by microscopical inspection of H&E, LFB-PAS and PLP-stained slices using an Olympus BX50^®^ microscope by two readers (VE, SH). Non-lesional normal appearing white matter (NAWM) was identified by well-preserved myelin in both LFB-PAS and PLP stainings. Diffusely abnormal white matter (DAWM) had ill-defined areas of LFB-PAS but no PLP reduction (28). Lesional WM included slowly expanding lesions (SEL), inactive lesions (InaL), shadow plaques (SdP) and ischemic lesions (IsL). Slowly expanding lesions had few LFBand PLPpositive myelin degradation products within phagocytes at the lesion borders, while their centers were almost completely devoid of myelin in the PLP and LFB stainings. Macrophages in the slowly expanding lesion cores were LFB-negative and frequently harbored PAS-positive material. Inactive lesions occasionally showed PAS-positive macrophages, but no LFB- or PLP-positive myelin remnants within phagocytes anywhere. Shadow plaques displayed sharp borders of LFB-myelin reduction, resulting from thinner remyelinating sheaths, and evenly distributed intact myelin throughout the plaque. We further encountered circumscribed lesions of myelin reduction but not depletion, similar to shadow plaques. However, these lesions had ill-defined borders and increased LFB-intensity around blood vessels traversing the lesions, i.e. a non-homogenous LFB-intensity reduction within the lesion. Their coarse texture indicated pronounced widening of the extracellular spaces. These were labeled ischemic lesions. ROIs were manually outlined on the LFB-PAS images. Anatomical and pathological boundaries were generously spared. To account for small differences in slice matching and imperfect registration, the masks were adapted to optimally match the TBB and PLP scans and the registered T_2_-weighted MRI. CD68 stainings were qualitatively evaluated for the distinction between slowly expanding (CD68 rim of microglia/macrophages present) and inactive (no CD68 rim present) lesions. BCAS1 stainings were evaluated with a counting grid in NAWM, DAWM and centers of slowly expanding, inactive and remyelinated lesions. The evaluated area covered 1 mm^2^, i.e. four neighboring visual fields at 20x magnification. Only clearly and strongly BCAS1-positive glial cells were counted.

### Data and statistical analysis

ROIs were automatically matched using their voxel location and the classification assigned to each ROI. We applied bwconncomp, a region-detection algorithm in MATLAB, to correlate histological and MRI values in all ROIs. Gray-scale medians for each ROI were calculated in all MRI and histological images, representative of each ROI’s myelin state.

Differences between slowly expanding and inactive lesions were assessed by a two-sided rank sum test (5% significance level). Correlations were performed on a per-case basis to suppress the influence of varying post-mortem delays, formalin fixation and storage times. Weighted average within-case slopes, intercepts and p-values were obtained, with weights represented by the variance of the individual within-section slopes, intercepts and correlation coefficients, respectively. Average correlation coefficients were determined by transforming the r-values using Fisher’s z-transformation and subsequently computing the hyperbolic tangent of the average of the transformed r-values. Finally, a linear mixed effects regression model was used in R to determine the standardized correlation coefficients as a measure of relative importance of LFB, PLP, TBB and storage time in predicting MWF and MTR (lm.beta package) (29). The data offset was modelled with two random factors that accounted for inter- and intra-subject variability. Collinearity between predictors was evaluated using the vif function (30). BCAS1 count differences were tested using an ANCOVA. ROI was used as fixed factor and slice ID as random factor to correct for multiple data points derived from the same cases.

## Results

Matching MRI and histology data, we obtained 27 section planes from the seven MS cases. GraSE MWF data of Case 5 were excluded due to unexpectedly low contrast, presumably related to changes in acquisition. One section from Case 6 was also excluded due to unreliable GraSE MWF signal. All cases showed pathological alterations typical of progressive MS, including DAWM, slowly expanding and inactive lesions as well as remyelinated shadow plaques. Classical active lesions were not observed. Major confounding pathology was noted in Case 7, which harboured ischemic lesions together with severely atherosclerotic large meningeal arteries. Only lesions of either clear ischemic or MS origin were analyzed. In total, 610 ROIs were outlined (median size 5.69 mm^2^, range: 0.14 mm^2^– 32.09 mm^2^). Figure 2 exemplifies matched MRI and histology with corresponding ROIs from four MS cases.

**Fig. 2.**
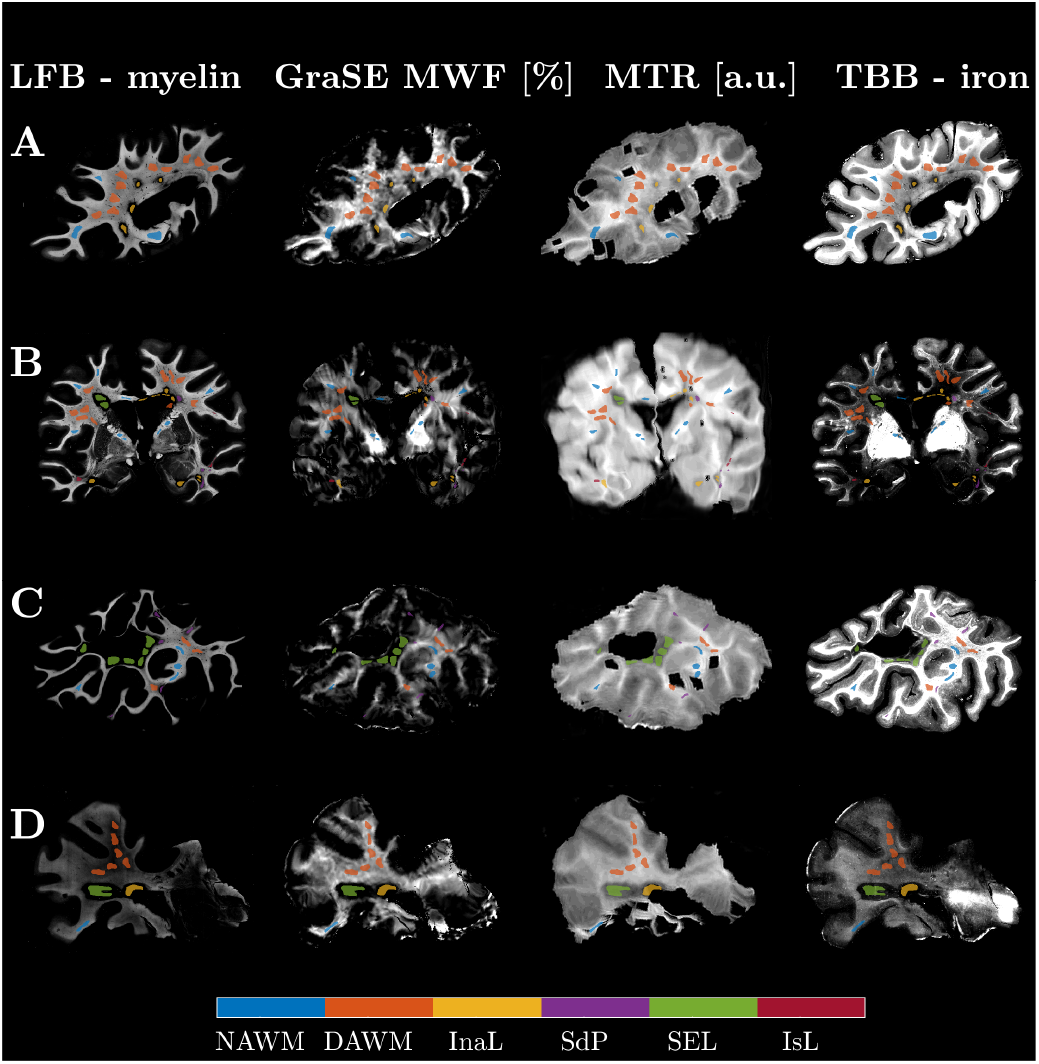
Qualitative comparison of the histology and co-registered MRI data for four sections from four cases. (A: Case 4, B: Case 7, C: Case 1, D: Case 2). The histological images were contrast-inverted to visually match the MRI contrasts so that lower intensities represent lower myelin or iron concentrations. Co-registration achieved good matching of the MRI to the LFB stains, which were chosen as histology reference. ROIs were centered in representative regions, yielding good local correspondence of ROIs on visual inspection. The ROIs were drawn with a gap to avoid partial volume effects at anatomical or lesion borders. (Acronyms: normal appearing WM (NAWM); diffusely abnormal WM (DAWM); inactive lesions (InaL); shadow plaques (SdP); slowly expanding lesions (SEL); ischemic lesions (IsL))

Histological stainings of one section of Case 7 are exemplified in Figure 3. NAWM was predominantly found in subcortical WM areas, since the progressive MS cases displayed typical widespread diffuse myelin injury in deep WM areas (Fig. 3g, black arrows). Inactive, but not slowly expanding lesions, harboured scattered thin myelin sheaths in their centers (compare Fig. 3i & Fig. 3j). BCAS1 staining (Fig. 3h) showed the highest cell densities in peri-plaque WM close to the rim of slowly expanding lesions (Fig. 3k), but on average hardly any positive cells in the lesion cores. A subset of inactive lesions, however, displayed clustered positive cells in the lesion center (Fig. 3l). Upon morphological evaluation of BCAS1+ cells, we noted that one portion of positive cells had strong cytoplasmic immunoreactivity, and either absent or very short and dystrophic-appearing processes. This type of BCAS1+ cell morphology was found in NAWM, DAWM and was most prominent in periplaque WM close to slowly expanding lesion (Fig. 3k). Conversely, BCAS1+ cells in inactive and remyelinated lesion cores had multiple, partly parallel processes (Fig. 3l). Ischemic lesions were reminiscent of shadow plaques regarding their myelin densities but differed morphologically. Shadow plaques were characterized by a sharp border and homogeneously thin myelin sheaths in the plaque area. By contrast, ischemic lesions showed a gradient of myelin intensity, including ill-defined borders, as well as a coarse lesion tissue texture with loss of whole myelin fibers, but preservation of normal myelin sheath thickness in the remaining fibers.

**Fig. 3.**
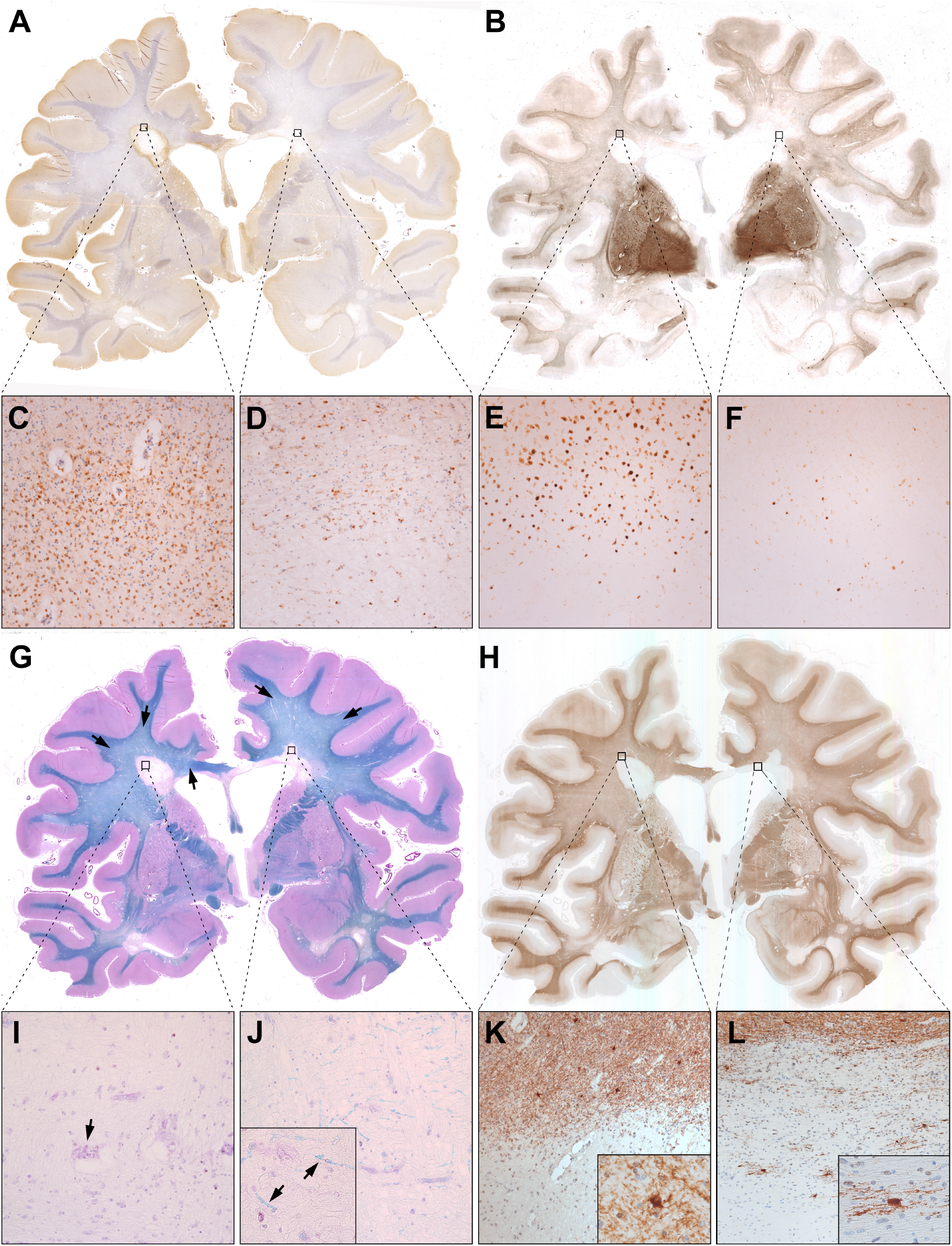
Histological sections of a double-hemispheric coronal slice level of Case 7. (A) CD68 immunohistochemistry revealed accumulated activated microglia and macrophages at the edge of a slowly expanding periventricular lesion (left black square, enlarged in C), but not at the inactive periventricular lesion edge (right black square, enlarged in D). (B) Similarly, TBB staining showed iron accumulation at the slowly expanding (E), but not at the inactive lesions edge (F). Both lesions had iron loss in their centers. In WM, iron staining was highest subcortically, gradually decreased in DAWM and towards lesions, and was lowest within lesion centers. (G) LFB-PAS myelin staining visually distinguished areas of NAWM (mainly subcortically and in the capsula interna), DAWM and MS lesions. Black arrows indicate the ill-defined DAWM, characterized by reduced myelin intensity. Both periventricular MS lesions (enlarged in I and J) showed myelin loss. Notably, scattered thin myelin sheaths were present within some inactive lesion centers (J, high magnification inset with arrows), but never in slowly expanding lesions (I). Occasional macrophages with PAS-positive cytoplasmic inclusions, indicative of remote demyelination, were found. (H) BCAS1 immunohistochemistry revealed moderate immunoreactivity in myelin, rendering exquisite histological myelin contrast. Strongly BCAS1-positive glial cells without processes and dystrophic morphology accumulated in areas of WM damage, such as DAWM (see quantitative data) and WM abutting slowly expanding lesion edges (K). Process-bearing actively remyelinating oligodendrocytes could be found exclusively in few inactive lesions (L) and shadow plaques.

### Comparison of MRI-based myelin estimates

Fig. 4a displays the regression between GraSE and CPMG MWF (R^2^ = 0.862, slope = 0.858 %/%, intercept = -0.42% MWF), which demonstrates excellent agreement of these two myelin water scans. However, the regression of CPMG MWF and MTR was insignificant (Fig. 4c, R^2^ *<* 0.01). Note that CPMG data were only acquired in samples with storage times longer than 9 years. The MTR – GraSE MWF relationship was modelled by two different, storage time dependent regression lines (Fig. 4b). GraSE MWF and MTR correlated significantly in tissue blocks with storage times of *<* 1 year, (R^2^ = 0.436), but not in those with extended storage times (R^2^ < 0.01). Notably, the regression intercepts in all correlations with MTR were different from zero. In cases stored *<* 1 year, the intercept for GraSE MWF and MTR was 19.3 a.u.. In cases with extended storage, the intercepts were 14.2 a.u. and 14.5 a.u., when comparing GraSE MWF and CPMG MWF with MTR, respectively.

**Fig. 4.**
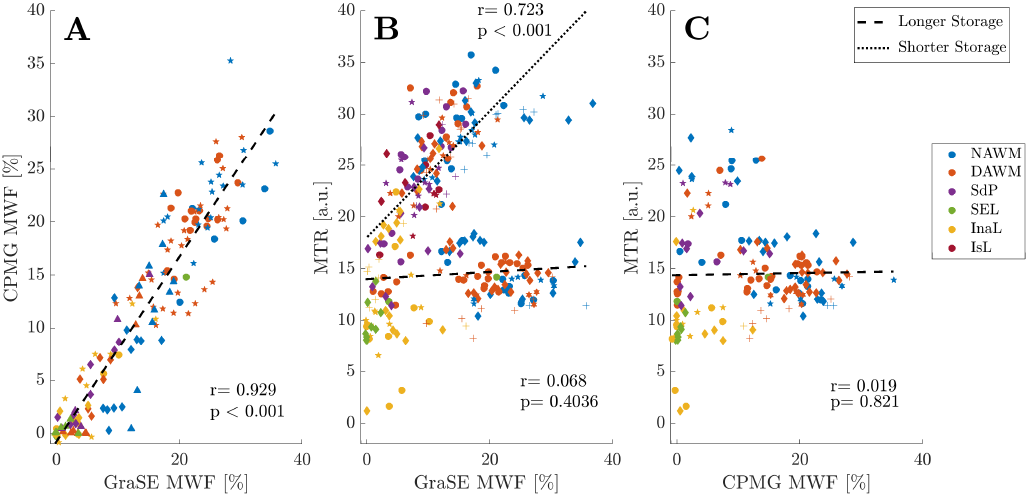
Correlation of all MRI metrics. Both MWF estimates showed excellent agreement with a minimal intercept (A), whereas correlations between MWF and MTR depended on storage times (B,C). Correlations between MWF and MTR were significant in tissues with storage times of *<* 1 year (dotted line) and were nonsignificant for tissues with extended storage time *>* 9 years (dashed line). Note that CPMG data were only available for samples with extended storage time (C). Symbols indicate different cases; colors distinguish different tissue types. (Acronyms: normal appearing WM (NAWM); diffusely abnormal WM (DAWM); inactive lesions (InaL); shadow plaques (SdP); slowly expanding lesions (SEL); ischemic lesions (IsL))

### Histological and MRI-based myelin measurements in different ROIs

Figure 5 displays the stratification of histological optical densities and MRI data according to ROI. WM LFB (Fig. 5a) and PLP (Fig. 5c) optical densities were highest in NAWM, followed by DAWM, shadow plaques and inactive lesions, and were lowest in slowly expanding lesions. LFB and PLP densities in ischemic lesions resembled those of shadow plaques. In contrast to LFB, PLP hardly differed between NAWM and DAWM. Iron was lower in DAWM compared to NAWM, and markedly depleted in slowly expanding and inactive lesions. In shadow plaques, iron densities ranged between the densities of DAWM and demyelinated lesions. Few high NAWM iron densities were derived from Case 1.

**Fig. 5.**
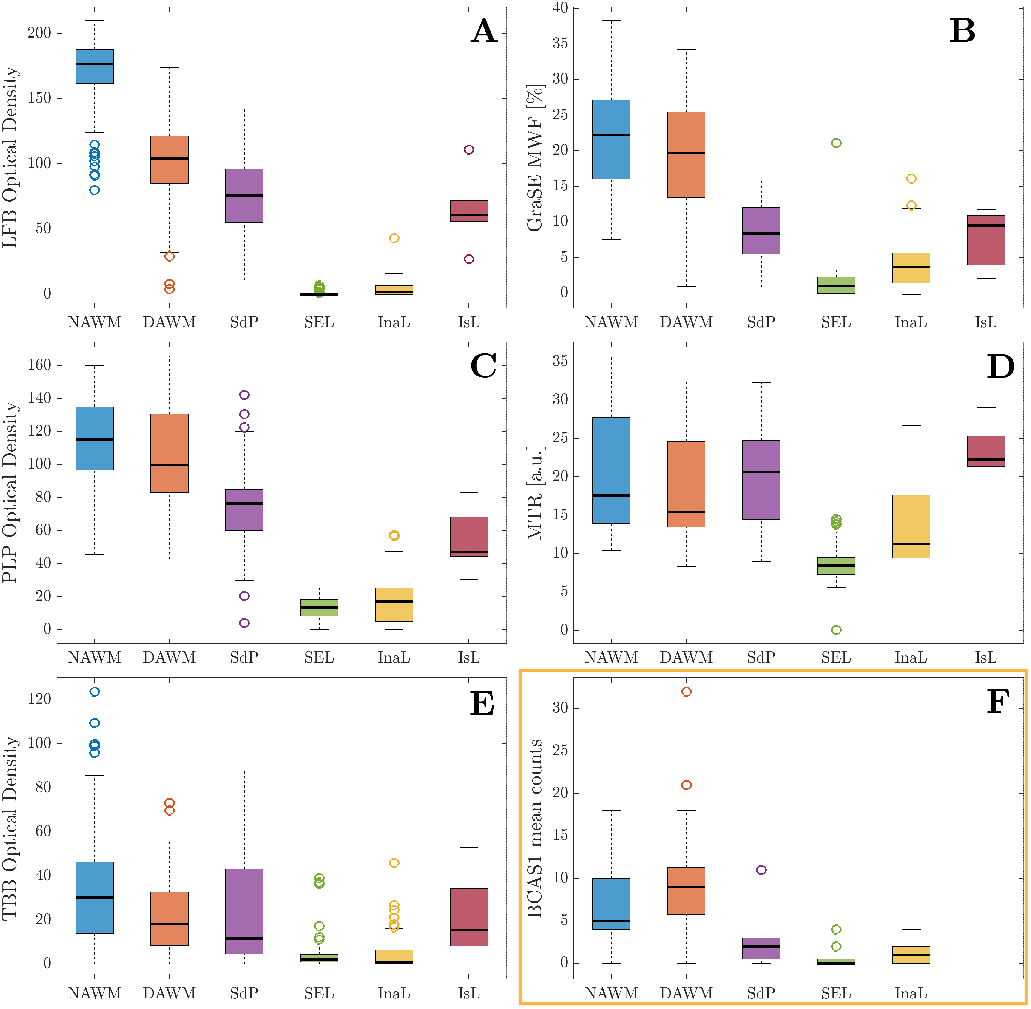
Comparison of quantitative MRI values and optical densities across regions of interest. Panels on the left display the histological density measurements (LFB (A), PLP (C), TBB (E)), panels on the right the MRI myelin measurements (GraSE MWF (B), MTR (D)). Panel F displays the separately obtained BCAS1 counts. LFB and PLP differences between ROIs, including in lesions, were well reflected in both MWF measurements. By contrast, MTR changes appeared more closely linked to the TBB staining intensities, and MTR values overlapped between regions. Note that the data collection for CPMG and MTR were incomplete, thus not all ROIs are present in the MTR plot. (Acronyms: normal appearing WM (NAWM); diffusely abnormal WM (DAWM); inactive lesions (InaL); shadow plaques (SdP); slowly expanding lesions (SEL); ischemic lesions (IsL))

Of all MRI metrics (Figs. 5b and 5d, CPMG data not shown), relative differences in GraSE MWF between ROIs most closely matched the LFB and PLP intensity distributions (Figs. 5a and 5c). All myelin MRI metrics were reduced in DAWM compared to NAWM, though less so than LFB. MWF was lower in all WM lesions than in NAWM or DAWM. In shadow plaques and ischemic lesions, MWF was between slowly expanding lesions and DAWM, best reflecting LFB and PLP relations. By contrast, MTR in shadow plaques exceeded NAWM MTR. All three MRI measures yielded higher values in inactive compared to slowly expanding lesions, sensitively reflecting histological myelin data. LFB (p *<* 0.001), GraSE MWF (p = 0.016) and MTR (p *<* 0.001) were all significantly higher in inactive compared to slowly expanding lesions, also after Bonferroni correction. BCAS1 counts, indicative of the number of actively remyelinating oligodendrocytes, were generally in line with the observations from LFB and MWF. The number of BCAS1+ cells in lesion areas was generally lower than counts in NAWM and DAWM. Slowly expanding lesions were found to have, on average, slightly lower BCAS1+ counts than inactive lesions and shadow plaques, although the difference was statistically not significant (p = 0.987). Significantly more BCAS1-positive cells were found in damaged DAWM compared to NAWM (p *<* 0.001).

Per-case correlations and average correlation coefficients (data not shown) between the MRI metrics and stainings reflect the findings of Figs. 4 and 5, i.e. strong correlations between myelin MRI metrics and myelin stainings, albeit with a nonzero intercept in correlations with MTR. Exemplarly, Figure 6 presents the GraSE MWF vs. LFB correlations. Note that sections containing only NAWM or DAWM, i.e. no lesions, exhibited typically insignificant, flat correlations and that MWF and MTR values varied between cases.

**Fig. 6.**
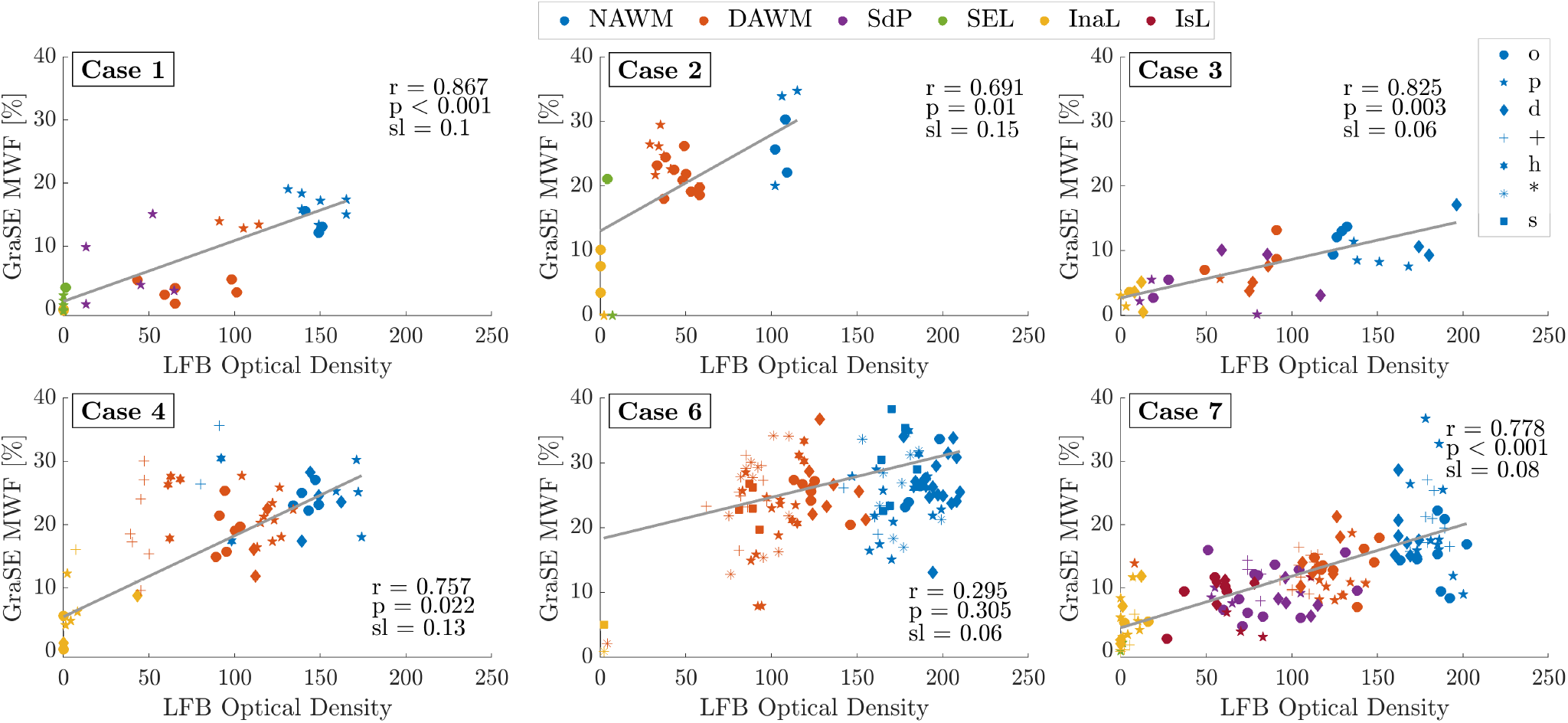
Within-case correlations between LFB and GraSE MWF across all ROIs. In order to suppress between-section deviations, all correlations shown are weighted averages of the individual within-section correlations. Different symbols distinguish data from different sections of a case. Different colors indicate different ROI types. The slopes of the average correlations ranged from 0.06 to 0.15 % MWF per optical density unit. All correlations, except for Case 6, were significant. Case 5 is not shown as the GraSE data were deemed unreliable for analysis. (Acronyms: normal appearing WM (NAWM); diffusely abnormal WM (DAWM); inactive lesions (InaL); shadow plaques (SdP); slowly expanding lesions (SEL); ischemic lesions (IsL))

Despite moderate to strong correlation coefficients across different tissue classes, within tissue class correlation coefficients were substantially smaller. To determine the extent to which LFB, PLP and TBB, in combination, predict the MRI metrics within specific tissue classes, we applied a linear mixed effects model. Data from all cases were included, and time since fixation was added as a fixed effect to account for differences in the MRI metrics related to storage time. Table 2 summarizes the significant contributors that were determined in the regression model. BCAS1 data were not included in the mixed effects model as they were separately obtained and analyzed.

**Table 2.**
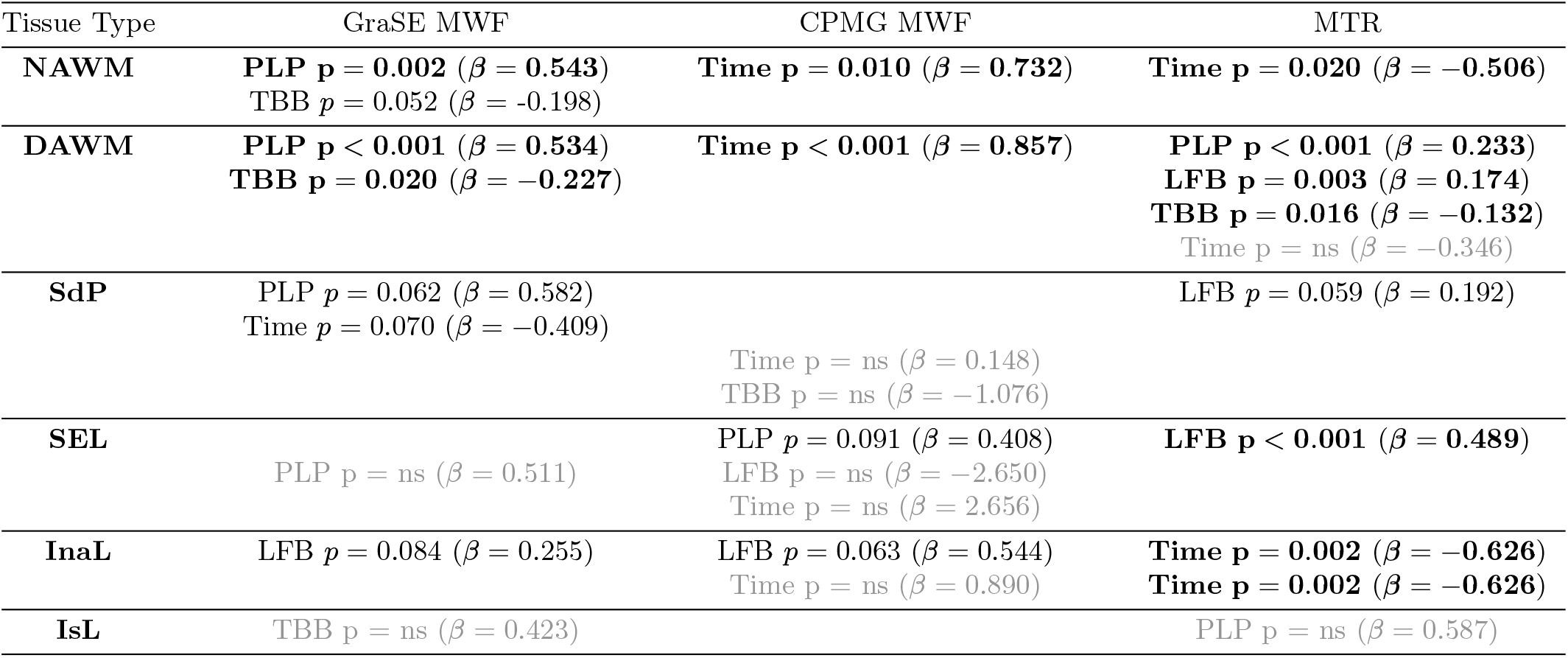
Linear mixed effects model ANOVA predictors of MR myelin measurements. Unadjusted p-values are reported together with their relative importance coefficients. Values reported in bold font are p *<* 0.05, non-bolded values reflect p-values between 0.05 and 0.1. If no significant factor was reported by the ANOVA (ns) or the largest ***β***-coefficient did not correspond to the significant entry, the highest importance factor is listed in gray in the second half of the respective cell. (Acronyms: normal appearing WM (NAWM); diffusely abnormal WM (DAWM); inactive lesions (InaL); shadow plaques (SdP); slowly expanding lesions (SEL); ischemic lesions (IsL))

| Tissue Type | GraSE MWF | CPMG MWF | MTR |
| --- | --- | --- | --- |
| NAWM | PLP **p = 0.002** ($\beta = 0.543$)<br>TBB *p* = 0.052 ($\beta$ = -0.198) | Time **p = 0.010** ($\beta = 0.732$) | Time **p = 0.020** ($\beta$ = -0.506) |
| DAWM | PLP **p < 0.001** ($\beta = 0.534$)<br>TBB **p = 0.020** ($\beta$ = -0.227) | Time **p < 0.001** ($\beta = 0.857$) | PLP **p < 0.001** ($\beta = 0.233$)<br>LFB **p = 0.003** ($\beta = 0.174$)<br>TBB **p = 0.016** ($\beta$ = -0.132)<br>Time *p* = ns ($\beta$ = -0.346) |
| SdP | PLP *p* = 0.062 ($\beta$ = 0.582)<br>Time *p* = 0.070 ($\beta$ = -0.409) | Time *p* = ns ($\beta$ = 0.148)<br>TBB *p* = ns ($\beta$ = -1.076) | LFB *p* = 0.059 ($\beta$ = 0.192) |
| SEL | PLP *p* = ns ($\beta$ = 0.511) | PLP *p* = 0.091 ($\beta$ = 0.408)<br>LFB *p* = ns ($\beta$ = -2.650)<br>Time *p* = ns ($\beta$ = 2.656) | LFB **p < 0.001** ($\beta = 0.489$) |
| InaL | LFB *p* = 0.084 ($\beta$ = 0.255) | LFB *p* = 0.063 ($\beta$ = 0.544)<br>Time *p* = ns ($\beta$ = 0.890) | Time **p = 0.002** ($\beta$ = -0.626)<br>Time **p = 0.002** ($\beta$ = -0.626) |
| IsL | TBB *p* = ns ($\beta$ = 0.423) | | PLP *p* = ns ($\beta$ = 0.587) |

PLP significantly explained variations in NAWM and DAWM GraSE MWF, while CPMG MWF and MTR were strongly influenced by tissue storage time. TBB significantly related to DAWM GraSE MWF and MTR. Similar to NAWM, MRI myelin values in shadow plaques were predicted by PLP, storage time and in part LFB. In line with the above-described differences between inactive and slowly expanding lesions, the mixed effects model revealed some association between LFB and MWF in inactive, but not in slowly expanding lesions. The latter only showed a dependency for MTR on LFB. Standardized regression coefficients, i.e. the amount of change that a variation in each of the predictor variable yields in the MRI-measured myelin values, are provided together with the p-values in Table 2.

## Discussion

We assessed the ability of three myelin-sensitive MRI techniques to differentiate various degrees of myelin damage, recovery and lesional activity in a larger sample of progressive MS cases. Using histochemical and immunohistochemical stainings for myelin, we showed low levels of myelin in completely inactive lesions. We suggest that this myelin signal may represent remyelination occurring in chronic MS lesions without microglia or iron rims. These low myelin levels were conspicuous in the high-magnification of the histological stainings and captured by all investigated myelin-sensitive MRI techniques and in part corroborated by BCAS1 stainings. Importantly, our data showed that BCAS1-positivity is not exclusively a marker for remyelinating oligodendrocytes, but that it also marks degenerating glial cells in DAWM and in the vicinity of the chronic active lesion edges. Detecting and following subtle remyelination in vivo in chronic lesions provides an additional avenue for assessing repair in MS and may be a relevant consideration for trials of remyelinating therapies, which typically are believed to be most efficacious in younger MS lesions.

### Comparison of MWF, MTR and histopathological metrics

The excellent agreement of CPMG and GraSE MWF (R^2^ = 0.862) corroborates in vivo comparisons (15). We attributed the two different relationships of MWF with MTR to storage time. MTR is known to decrease with formaldehyde fixation (31), but longer-term pH changes may also affect MTR similar to other off-resonance techniques (32). NAWM and DAWM showed the most pronounced storage-related MTR shifts of up to 20 a.u., much more than the difference in intercept. Between cases, acquisition differences and differences in iron content affect MTR. Increased TR is known to result in MTR reductions (33), while strong WM iron depletion significantly increases MTR (22, 24, 25). More work is warranted to understand the dependency of MTR on the storage duration after formalin fixation.

Independently, non-zero regression intercepts between MTR and LFB, PLP and the other two MRI myelin measures indicate that MTR reflects not only myelin lipids and proteins but also other tissue characteristics, despite being positively correlated with high R^2^. Although MTR is widely used to detect deand remyelination (34), other non-aqueous tissue components, such as axonal and glial cell membranes, soluble proteins in the extracellular space, and astrocytic and axonal densities also play a role (35). Inflammation and edema further confound the specificity of MTR for myelin (35, 36), while MWF remains relatively unchanged (37, 38). Note the large overlap in MTR across different tissue types and the relatively high MTR in slowly expanding and inactive lesions, resulting from the non-zero intercept.

Limitations of earlier MWI approaches (5, 16), i.e. small field-of-view and very long acquisition times, have been overcome by introducing the 3D GraSE sequence in MWI (15). However, the now widely used GraSE-MWI approach has never been histopathologically validated. Previous MWF histopathological comparison focused on LFB intensities (8) and recently,electron microscopy validated CPMG-MWI in rat spinal cord samples (9). We assessed myelin more broadly, including lipids as well as myelin proteins and iron to explain variations in MWF (28), with CD68 and BCAS1 for further validation. Gradient echoes in the GraSE sequence may make MWF more susceptible to static field inhomogeneities. However, in agreement with other work (24), we did not observe differences between GraSE and CPMG MWF, although TBB contributed to the prediction of GraSE MWF in some regions (Table 2). Note that iron was highly variable between cases in the present study, possibly due to reduced tissue iron concentrations in progressive cases with long disease duration (22).

Relative differences in MWF between tissues agreed well with the differences in LFB and PLP stainings, including in lesions. This demonstrates excellent sensitivity of MRI-based myelin imaging to the detection of different stages or degrees of deor remyelination, despite low lesional myelin concentrations. Integrating the different stainings in a multi-linear regression showed that both myelin lipid and protein concentrations are needed to predict variations within tissue classes, such as within DAWM or between different shadow plaques. Notably, LFB and PLP explained much greater variation in MWF than TBB.

### Slowly Expanding and Inactive Lesions

Differently from slowly expanding lesions (39), inactive lesions do not have a rim of degradation products, but both are typically described as relatively free of myelin containing macrophages, microglial activation, and inflammation in their lesion centers (40). By contrast, we observed non-zero MWF and MTR in inactive lesions and found few, thin myelin sheaths in LFB sections, not seen in slowly expanding lesions (Figure 3). LFB and both MWF and MTR showed consistently higher levels in inactive compared to slowly expanding lesions. Correspondingly, the statistical model detected, albeit insignificantly, that LFB influenced GraSE and CPMG MWF for inactive, but not for slowly expanding lesions.

Our findings are in line with post-mortem work showing insignificantly higher MTR and macromolecular proton fractions in inactive compared to slowly expanding lesions (35). Moreover, in vivo susceptibility and myelin imaging demonstrated that rim lesions have lower myelin values than inactive lesions (41). Most of these studies attributed the lower signal to the greater destructiveness of slowly enhancing lesions. Meanwhile, it has been shown that both slowly expanding and inactive lesions may contain BCAS1+ premyelinating oligodendrocytes, indicating the ability of both lesion types to remyelinate (19). It is believed that while the rim of activated microglia and macrophages exists, it may prevent lesional myelin recovery (39, 42). Our data suggest that inactive lesions, for which the rim has resolved, start to recover some thin myelin sheaths to a degree that the lipid signal can be detected on histological stainings and with myelin-sensitive MRI techniques.

Thus, it may be possible to capture varying degrees of late remyelination also in vivo using MRI. All these observations warrant further investigation to better characterize the difference between slowly expanding and inactive lesions.

Notably, our data showed that BCAS1+ also stains degenerating glial cells. This is supported by the BCAS1+ cell morphology in areas of DAWM and periplaque WM, where cells had either no or short and twisted dystrophic processes. By contrast, in few areas within inactive or partly remyelinated lesions many BCAS1+ cells had extensive branches and formed processes to multiple internodes, yielding a morphology reminiscent of that described for PLP-positive myelinating oligodendrocytes (43). However, this was only found in few lesions and might relate to the transient nature of remyelination.

### Normal appearing and diffusely abnormal WM & Shadow Plaques and Ischemic Lesions

The lower LFB but similar PLP signal between DAWM and NAWM (Figure 5) agrees with the lipid abnormality characteristic of DAWM (28). Interestingly, our mixed effects model suggested that more subtle differences within NAWM and DAWM GraSE MWF relate to differing PLP-concentrations. TBB also played a role in distinguishing subtle differences between NAWM and DAWM regions, in line with the importance of iron for myelin production in oligodendrocytes and previously described iron loss in DAWM (44). Given the widespread diffuse injury, NAWM ROIs were confined to subcortical regions. These tissue block areas are more likely to be affected by misregistration errors, possibly explaining why the MRI-measured NAWM-DAWM difference was smaller than expected from their LFB intensities. CPMG and MTR were predominantly related to storage time or a combination of predicting factors.

Shadow plaques and ischemic lesions were well reflected in GraSE MWF, but both lesion types had higher MTR than NAWM and DAWM. This may again relate to the inverse relationship between MTR and iron concentrations (23, 25). Note that our shadow plaques yielded non-zero TBB staining, although they have been generally found to remyelinate without iron recovery (22). Regardless, myelin-related regression coefficients for GraSE MWF in shadow plaques were noticeably larger than the TBB coefficients, indicating that iron plays a minor role in the estimation of MWF in shadow plaques. No specific histopathological factor predicted MWF or MTR of ischemic lesions.

### Study strengths and limitations

We tested how myelin lipids, proteins and iron contribute to MWF and MTR. Our data provide the first validation of the use of the GraSE sequence for MWI. We demonstrated the sensitivity of these myelin imaging techniques on a case-by-case basis. Across cases, we utilized mixed effects modeling to provide a combined assessment of the role of lipids, proteins, and iron to the MRI measures, rather than assessing them in isolation. We did not consider axonal density, which is relevant for MTR (35), due to its strong collinearity with myelin. Assessing gliosis and inflammation, however, may help to shed further light on the observed lesional differences. BCAS1 staining was performed separately to support our initial findings. Dedicated sample selection to investigate specific tissue classes or class differences would be advantageous but was beyond the scope of the work presented here.

Our investigation included MTR (6), rather than quantitative magnetization transfer (35), as MTR is more frequently used, both in clinical trials and research. Assessing other techniques, e.g. 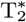-based myelin mapping (7, 45), may be of interest in the future. Although other MRI techniques may have advantages over traditional MWI acquisitions, including potentially faster acquisition times (6), higher spatial resolution (46), improved signal-to-noise ratio and lower specific absorption rates, they are also known to be influenced by axonal water (12), magnetization transfer effects (11, 36, 47) or to suffer phase errors due to field inhomogeneities and physiological noise (48, 49). Current MWI approaches (14, 21) also have the advantage of being model-free, and new developments in MWI may alleviate limitations in scan time and spatial resolution (50).

## Conclusions

Both MWI strategies, the original CPMG and the faster GraSE acquisition, as well as MTR reflected the histologically assessed myelin concentrations across tissues and allowed to detect subtle within-tissue class differences. Notably, the range of MWF and MTR values observed in inactive lesions appeared to reflect variable degrees of late, slowly progressing remyelination.

### Data Availability Statement

Data are available from the corresponding author upon reasonable request.

## Data Availability

Data are available from the corresponding author upon reasonable request.

## ACKNOWLEDGMENTS

The authors wish to thank Marianne Leisser for supporting the histological analysis at the Medical University of Vienna, and Professor Hans Lassmann for allowing them to use tissue from the multiple sclerosis brain bank for this work. We wish to acknowledge the continued research support by Philips Healthcare. This study was supported by the Research Methodology Grant from the BC Children’s Hospital Research Institute (former CFRI) and funding from the National Multiple Sclerosis Society (RG-1507-05301). VW was supported by a graduate student award from the Multiple Sclerosis Society of Canada (EGID 2002). VE was supported by funding from the National Multiple Sclerosis Society (RG-1507-05301). EHT was supported by Consejo Nacional de Ciencia y Tecnologia (237961). We are grateful for additional support from NSERC (402039-2011, 2016-05371), CIHR (RN382474-418628) and the Milan and Maureen Ilich Foundation. AR is supported by Canada Research Chairs (950-230363).

